# Læmple: A Benchmarking Framework for Virus Lineage Deconvolution Tools for SARS-CoV-2 from Wastewater

**DOI:** 10.64898/2026.09.02.26362081

**Authors:** Anna Schedl, Andreas Bergthaler, Fabian Amman

## Abstract

1

**Background:** Correct and accurate deconvolution of SARS-CoV-2 lineages from wastewater sequencing data is a challenging task, given the intricacy of wastewater amplicon-sequencing data and the ever-growing complexity of the lineage classification. Existing benchmarking studies made use of artificial spike-in compositions, thereby falling short of reflecting the prevailing complexity of wastewater samples.

**Results:** We present a modular, expandable, simulation-based benchmarking framework as a reproducible Snakemake workflow, named Læmple, to evaluate the performance of virus lineage deconvolution tools. Using in silico simulated data sets of varying complexity and sequencing quality, we demonstrate its utility by evaluating seven publicly available tools, based on their precision, sensitivity, and reproducibility. Freyja showed robust sensitivity and consistent performance across diverse data set complexities, alongside user-friendly installation and documentation, while VaQuERo demonstrated the highest precision.

**Conclusions:** Our results reveal substantial variation in tool performance across conditions, emphasizing the need to benchmark with diverse and complex scenarios. This framework enables informed tool selection for researchers and public health agencies and allows developers to stress-test their tools during development and maintenance, i.e., updating the lineage definition for newly emerging virus lineages. To this end, Læmple was designed in a modular fashion for customization and future expansion to support ongoing software development across all stages of the application life-cycle management.

## 2 Background

The COVID-19 pandemic has highlighted the importance of genomic pathogen surveillance for public health. While individual case monitoring remains the gold standard for genomic surveillance, especially to link viral variants to clinical outcomes [1], it is resource-intensive [2] and prone to sampling bias [3]. Wastewater-based surveillance (WBS) offers a cost-effective and non-invasive alternative, capturing community-wide viral trends while avoiding individual testing limitations [4, 5]. Sequencing of the virus matter from wastewater additionally allows one to deduce the genetic make-up of the circulating virus in the population. The conglomerated nature of wastewater samples, and the general error-proneness of wastewater sequencing data, makes deconvoluting the presence and abundance of different virus lineages in such a sample a formidable task. To this effect, multiple bioinformatics tools for estimating SARS-CoV-2 lineage prevalences from complex wastewater samples have been developed [6, 7, 8, 9, 10, 11, 12, 13, 14, 15, 16, 17] and are still being developed [18, 19].

A robust lineage deconvolution is a prerequisite for downstream modeling efforts. Thereby, different tasks, such as the detection of a new variant of interest, or the characterization of lineages to infer their individual growth advantages, require different balances between sensitivity and specificity. In turn, sensitivity and specificity are an intricate product of the applied algorithm, its guiding parameters, the underlying variant definition, and last but not least, the input sample’s lineage composition complexity. The former two are somewhat static problems, for which researchers and public health practitioners need to evaluate the currently available spectrum of deconvolution tools once and settle on the most suitable set-up. The latter two factors are more obfuscated since they can steadily shift while the progression of the pathogen’s evolution requires new lineage definitions and the set of co-circulating lineages is ever-changing. Consequentially, a low-threshold but still thorough evaluation of the available deconvolution tools’ performance is pivotal during software development and maintenance, wastewater monitoring program design, and its execution.

Addressing this challenge requires benchmarking frameworks which enable users to systematically evaluate tool performance under conditions relevant to their specific use cases. While several benchmarking studies have evaluated different combinations of tools using simulated data, spike-in experiments, or real sequencing data [20, 21, 14, 22, 23], these efforts typically present fixed benchmarking setups tailored to specific tools or scenarios. In contrast, rather than introducing another tool-specific comparison, we present a general benchmarking workflow that enables users to perform their own standardized and reproducible evaluations across diverse data types and use cases.

A central design choice of this workflow is the use of simulated data. Simulation not only lowers the barrier for users to generate custom benchmarking data sets tailored to their specific questions, but also allows precise control over lineage composition, abundance, and evolutionary relationships. Importantly, this choice also addresses a key limitation of many existing spike-in benchmarking experiments, which often rely on simplified mixtures and therefore fail to capture the complexity observed in real-world lineage dynamics.

In contrast, studies employing in silico simulated data sets are more flexible in this regard [23], especially since artifacts typically seen in wastewater can be reflected during the simulation process [24]. It was already recognized that these first studies do not reflect the needed breadth to cover the many scenarios relevant to assess the performance of individual deconvolution tools [25].

To address this, we have developed a flexible and adaptable in silico benchmarking framework for repeated evaluation of virus lineage deconvolution tools in a multitude of different scenarios. Building on SWAMPy, a dedicated sequencing data simulation tool for wastewater data [24], our workflow supports the generation of well-characterized, in silico simulated data sets with arbitrary lineage mixture complexity and variable sequencing quality, allowing for robust and flexible performance assessments. By integrating data simulation, tool execution, and performance metric evaluation within a single Snakemake workflow, the framework facilitates direct comparisons between tools, versions, parameters, and scenarios, thereby supporting informed decision-making for adopters and tool developers. As a demonstration of the framework’s utility, we applied it to seven publicly available deconvolution tools and provide a detailed performance comparison. However, the primary contribution is the benchmarking framework itself, which is designed to support ongoing tool evaluation, development, and informed decision-making, both in academic and public health contexts.

## 3 Workflow description

The presented workflow is implemented as a Snakemake workflow [26]. It consists of four main steps: (i) customized input sequencing data simulation, (ii) mutation calling, (iii) sub-modules for lineage deconvolution by individual tools, and (iv) integrative performance analysis to compare against simulated ground truth. Thereby, it simulates sequencing data reflecting lineage abundance timecourse trajectories of arbitrary complexity, analyzes the data with pre-selected tools already available as sub-modules and produces informative comparisons between the assessed tools’ performance. Future users can easily adapt the workflow by providing new analysis modules for their own deconvolution tools, or by adapting an existing one to compare different parameters or versions of a tool in different scenarios.

### 3.1 Input data simulation

To systematically evaluate lineage deconvolution methods, we developed a flexible simulation pipeline capable of generating synthetic SARS-CoV-2 sequencing data sets with time-resolved lineage abundances. For empirical realism, a lineage-resolved pandemic progression, consisting of relative lineage abundances per time point, is introduced. This can be a user-specified input, with arbitrary complexity, or it will be constructed from a list of specified lineages, their date of emergence and their relative growth advantage. In the latter case, lineage dynamics follow a logistic growth model, under the assumption that newly introduced variants will gradually overtake earlier ones in prevalence. Subsequently, a user-defined number of samples is drawn from the underlying time-course. Sampling strategies include evenly spaced intervals, random time-point sampling, or exhaustive sampling, thereby providing flexibility for simulating different study designs or sequencing campaigns. For each of the drawn samples, a simulated sequencing data set is generated using SWAMPy [24]. Besides variant abundances from the previous step, SWAMPy requires virus lineage reference genomes, a corresponding amplicon primer scheme, and a quality parameter as input to produce simulated NGS reads.

For our showcase analysis, all NGS data were simulated using the ARTIC Network V4 Primers. The specified quality parameter controls the uniformity of reads per amplicon; therefore it enables the adjustment of the whole-genome coverage of the produced reads. SWAMPy mimics characteristics of real-world wastewater-derived sequencing reads by considering RNA degradation, PCR errors, and other library preparation artifacts, as empirically deduced from real wastewater experiments. This modified amplicon population is then used as input to simulate sequencing reads using ART, a multipurpose next-generation sequencing read simulator [27]. For our analysis, seven different quality settings are simulated, leading to whole-genome coverages between 4 % and 97 %. Additionally, the default error model for 150 bp paired-end reads of the Illumina MiSeq V3 Sequencing system, both for sequencing errors and base quality is applied. Other error profiles enabled by the underlying ART read simulator, are available, but limited to Illumina systems.

### 3.2 Mutation calling

Deconvolution tools differ in their input requirements. Some demand raw reads and provide internal functionality for the detection of nucleotide polymorphisms in the data. Others require already quantified nucleotide polymorphisms as input, typically in the form of a VCF file format. To account for this, a previously described variant calling approach [28] was implemented as a dedicated Snakemake module. In brief, overlapping regions of read pairs are corrected using bbmerge; version 39.01 [29], then mapped against the reference genome via BWA-MEM; version 0.7.17 [30]. All mapped reads are filtered, and primer sequences removed via iVar; version 1.4.2 [31], and subjected to the Viterbi and indelqual method of the LoFreq tool; version 2.1.5 [32], after which, due to occasional excessive regional read coverage and consequently exuberant run time, reads are down-sampled to a maximum of 10,000 read starts per genomic position, to streamline processing time. Afterwards, LoFreq and Bcftools; version 1.17 [33] are used to filter for variants with a minimum coverage of 75 reads, a minimum Phred-scaled calling quality of 90 and indels with an HRUN value of less than 4. Produced VCF files serve as input for subsequent lineage deconvolution tools, if appropriate.

### 3.3 Tool specific modules

Each deconvolution tool is integrated into its own module within the overall benchmarking workflow. These modules receive input data generated by the previously described mutation calling pipeline and execute the respective tool. Eventually, an additional formatting step ensures that all tool outputs are converted into a unified, standardized format compatible with the downstream performance analysis. For the purpose of the current workflow implementation, all tools are installed and parametrized using their recommended configurations according to their documentation. The modular structure enables straightforward tool removal from or insertion into the overall workflow, allowing for easy updates or expansion. New tools can be integrated by adding additional modules, making the framework highly adaptable for changing needs or different research scopes. A discussion of initially available tool-specific modules can be found below.

### 3.4 Performance Analyses

In a final module, the presented framework evaluates both qualitative and quantitative performance of lineage deconvolution tools, such as lineage detection and relative abundance estimation. Furthermore, reproducibility across technical replicates and the impact of low-quality sequencing input data is assessed. Finally, performance comparisons are visualized in expressive graphics, allowing users to quickly gain insights about relative performance differences. For an overview of the complete report see Supplemental File 1.

#### 3.4.1 Variant detection

Key metrics for measuring the reliability of lineage detection are True Positive Rate (TPR), also called recall or sensitivity; False Negative Rate (FNR) or miss rate; Positive Predictive Value (PPV) or precision, as well as the Jaccard index (JI). These metrics measure the overlap between predicted and simulated (true) lineage sets and are independent of True Negative values, which are dependent on the size of the reference set available to the tool and therefore most sensitive to user-predefined input parameters in particular the volume of the tool specific lineage search space. All key metrics are calculated for each experimental replicate and median results are reported.

The above performance metrics consider a binary true/false classification and neglect the severity of misclassifications. Erroneous detection of closely related lineages, with few distinct mutations, can be considered less grave than false calls from unrelated clades. To account for this, each false positive is further contextualized in relation to the most similar true positive lineage from the simulated ground truth. This similarity is quantified using the Jaccard distance, based on shared defining mutations between each lineage pair. Furthermore, using the cladogram-like nature of the lineage nomenclature [34, 35], false positives are classified either as linear descendants (ancestor-descendant pair) or as originating from different sibling clades. In case of a linear relationship, directionality is also assessed by determining if the false positive is either a descendant (child) or an ancestor (parent) of the closest true lineage to provide a deeper insight into the nature of the misclassification.

#### 3.4.2 Variant abundance estimation

Relative lineage abundances per sample are recorded and compared to expected values. If predicted abundances in a sample do not sum up to 100 %, the remaining portion is assigned to an “others” category. Lineages with predicted abundance below a user-defined threshold (default set to 2 %) are also grouped into “others”. This step helps declutter results from low-confidence predictions and increases ease of interpretation. To evaluate qualitative performance, root mean squared error (RMSE) is calculated per sample, reflecting deviation from the simulated (true) lineage abundances. For false negatives (i.e., undetected simulated lineages), the corresponding inferred abundance is set to zero. For comparability, RMSE is computed only across the simulated (true) lineages, excluding false positives. However, false positives may still indirectly affect results by lowering the predicted abundances of other lineages.

## 4 Example Analysis - Benchmarking

To demonstrate the utility of our benchmark framework, we applied it to publicly available deconvolution tools. In this example, we focused on how sequencing quality and lineage dynamic complexity influence tool performance.

### 4.1 Pre-installed Tools

We include a selection of available tools, chosen for their popularity as public health tools, diversity of their underlying methodology, and ease of installation. More detailed notes about user experience during installation can be found in the Supplemental Material. Furthermore, for two selected tools (Freyja and VaQuERo), modules implementing different versions and different reference data sets are included. This allows us to showcase the utility of our framework to track performance effects during software development and maintenance. In the following, the default included modules and their associated tools are briefly discussed.

#### 4.1.1 Freyja

Wastewater genomic analysis with Freyja is based on lineage determining mutational barcodes derived from the UShER global phylogenetic tree. It uses single-nucleotide variants (SNVs) to characterize virus diversity and identify lineages by their respective barcodes. Observed mutation frequency is weighted against transformed sequencing depth to adjust for large differences in depth across amplicons and down-weight genomic positions with low coverage. Haplotype frequency is then inferred from a constrained, weighted least absolute deviation problem. Again, this should make it robust for low-coverage sequencing samples, or samples with very uneven coverage distribution over the whole genome [13]. For our example workflow we compared three different versions with their respective default reference data sets and parameters (v1.4.3, v1.5.3, v2.0.0), which were released between May 2024 and July 2025.

#### 4.1.2 Lineage deComposition

Lineage deComposition (LCS) also uses a variant composition model built on pre-defined marker mutation sets, which are characteristic for a given lineage, but also incorporates a probability matrix for finding alternate mutations on any given position in any given lineage [17], derived from aligned, manually curated genomes designated to lineages corresponding to the Pango Lineage nomenclature [34, 35]. Estimates for proportions of each lineage in a sample are obtained using a maximization algorithm using counts of reads aligned to a reference sequence for each variant and the probability matrix. For the example workflow, LCS was installed via GitHub repository (commitID: cfc2f02), from 2024-08-27.

#### 4.1.3 Lollipop

Lollipop was specifically designed for deconvolution of SARS-CoV-2 lineages from wastewater samples [9] and forms part of V-pipe [36], a workflow designed for analysis of NGS data from viral pathogens used by the Swiss Federal Office of Public Health [12]. The tool aims to estimate relative abundances of genomic variants by using observed mutation frequencies, weighted according to a kernel function and deconvoluted according to lineage-defining mutation lists, while making predictions robust, even with lower-quality sequencing samples [9]. Lollipop v0.3.0 was installed via conda installation on 2023-05-29.

#### 4.1.4 QuaID

QuaID [15] uses a pre-generated multiple sequence alignment file from GISAID [37] to extract all mutations of any given lineage that appeared in at least 50 % of all genomes associated with that lineage and were considered quasi-unique for that lineage. Setting stricter thresholds for either parameter will lead to smaller sets of characteristic mutations with higher confidence, trading sensitivity for specificity.

In this study, we kept all thresholds at the default values, opting for a balanced approach. QuaID was installed on 2024-04-08 from the public GitHub repository (commitID: aec47270).

#### 4.1.5 VaQuERo

Similarly, detection of variants in VaQuERo (i.e., Variant quantification in sewage pipeline designed for robustness) is also based on predefined marker mutations, followed by a SIMPLEX regression model to quantify abundances of detected variants [6]. Mutation frequency is down-weighted by sequencing depth at the mutation position. In contrast to other tested tools, VaQuERo analyses time-course data by considering neighboring time-points with time-dependent weights in the regression model. For the example workflow, two different versions of VaQuERo v.2 were installed from the GitHub repository (commitID: 93e7db3 on 2023-08-21, and commitID: 24d9211 on 2025-08-16), as well as the newer version with the reference set from the older version 24d9211 OLDREF.

#### 4.1.6 Viral Lineage Quantification

Viral Lineage Quantification (VLQ) is a bioinformatic pipeline which uses kallisto [38], a tool originally designed for transcript quantification, for its lineage deconvolution step [7]. It maps sequencing reads to a reference set of transcriptional, or in this case, whole-genome virus sequences using a De Bruijn graph of k-mers to estimate read-reference sequence associations. Applicable to multiple data types, from whole-genome sequencing to spike-only amplicon sequencing, VLQ has shown a very low detection limit, under 1 % in simulated data, up to 10 % in real-world data [7]. VLQ was installed on 2023-05-29 from the public GitHub repository (commitID: d04ad36).

#### 4.1.7 Virpool

VirPool is using the probability of observed alternative nucleotides at any position in any lineage based on genomic sequences downloaded from the GISAID database aligned to a reference sequence, creating mutation profiles for each variant [11]. Adjusting for sequencing errors, the probability of observing specific reads originating from individual lineages is used to weigh each lineage present in a user-defined variant profile list. Using a softmax transformation, the VirPool algorithm optimizes for the highest likelihood of observed proportions of each lineage for a given set of sequencing reads and lineage profile list, making the results dependent on pre-selection of lineages to be included in the analysis. VirPool was installed on 2023-04-08 from the public GitHub repository (commit: 763c212).

## 5 Input Data Description

To evaluate tool performance across a range of conditions, we simulated three distinct scenarios designed to reflect increasing levels of complexity, by modulating the number and intricacy of cocirculating lineages. Each scenario spanned 365 time-points (representing one year) with samples taken every 14 days, for a total of 25 samples per year. At each time-point, sequencing reads were simulated based on the modeled lineage abundances and each sample was generated in triplicate using different random seeds to mimic technical replicates. To test robustness against sequencing quality,each triplicate was simulated at seven different coverage levels by adapting simulated genomic read coverage, resulting in 525 samples per data set in total. Two of the assessed scenarios followed a simplified logistic-growth model with 1 initial case and maximum of 10^6^ cases per newly introduced lineage and an exponential growth rate of 0.2. In all simulations, the SARS-CoV-2 Wuhan-01 wildtype sequence is included as a background signal at 2 % relative abundance. To examine the effect of lineage similarity on prediction accuracy, we looked at two extreme cases. One data set was simulated using very closely related lineages, namely different variants of the XBB.* sublineage, with an average of 88 % shared mutations (“Omicron time-course”). The other data set used more genetically distinct lineages, namely a selection of common variants of concern, with an average of 11 % overlapping mutations (“VOC time-course”). Finally, the third data set was designed to mimic real-world lineage dynamics. To this end, we used the weekly lineage frequencies as determined from case-based surveillance in Vienna/Austria during the period 2022-01-01 to 2023-01-01, based on 38,511 individual clinical samples (“Vienna time-course”). The collection time and the associated lineage annotation were derived from the GISAID database [37]. Sequences were filtered by submission date and location, aggregated weekly, and normalized for relative lineage abundance per week. Lineages present in fewer than 3 samples or less than 1 % frequency per week were excluded, as well as lineages which were detected in less than two weeks overall. In total, 92 lineages remain, mostly sub-lineages of BA.1 and BA.2. For post-prediction analysis, lineage-level abundance estimates were compared to simulated ground truth as described above.

## 6 Results

Our presented framework for the assessment of virus lineage deconvolution tools enables users to examine the performance of various tools in a streamlined manner. The raw and comprehensive overview of the results of the applied analysis is presented in Supplemental File 1. In the following section, notable outcomes and findings with respect to the assessed tools are highlighted, to showcase the versatility of application cases enabled by the Læmple framework.

### 6.1 Lineage Identification and Abundance Estimation Across Deconvolution Tools

To provide a comprehensive comparative overview of individual tool performances within each data set, we combined all results for the simulated data sets with different complexities, and plotted the Jaccard Index, summarizing identification accuracy, against the Root Mean Squared Error (RMSE), which reflects the accuracy of relative abundance estimation. Across all deconvolution tools evaluated, performance varied greatly in both lineage identification accuracy and abundance estimation error (Figure 1). Most tools clustered at low to moderate Jaccard Index values (0.04 - 0.34) with corresponding RMSE values between 0.12 and 0.23. VaQuERo versions showed similar performance to one another, with Jaccard Indices ~0.2 and RMSE values ~0.16. Freyja versions presented slightly higher Jaccard Indices (up to ~0.34) and moderate RMSE values (0.13-0.16). Two Freyja versions (v1.5.3 & v2.0.0) generally showed higher lineage identification accuracy and quantitative abundance prediction, while most other tools produced modest Jaccard scores with similar or slightly higher abundance errors.

**Figure 1:**
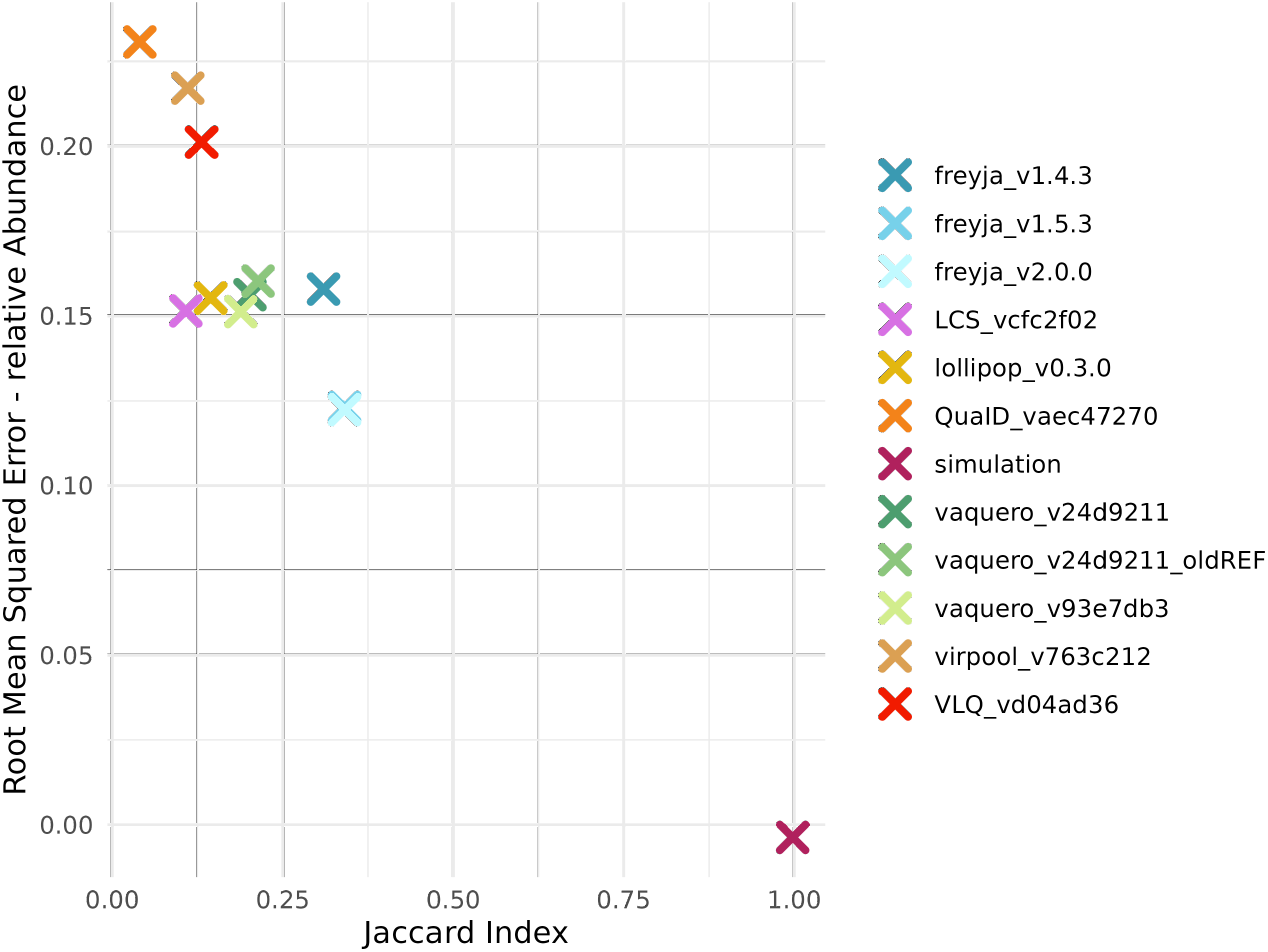
Comparison of lineage identification accuracy and abundance estimation error across multiple SARS-CoV-2 lineage deconvolution tools and versions aggregated across all simulated data sets. Each point represents a tool’s performance with the x-axis showing the Jaccard Index, for which higher values indicate higher accuracy in lineage identification. The y-axis shows Root Mean Squared Error (RMSE) of relative abundance estimates; lower values indicate better quantitative prediction performance. The mark *simulation* depicts the optimal performance.

### 6.2 Assessment of False-Positive patterns Across Deconvolution Tools

Across all tools and simulated experiments, false-positive lineage calls displayed distinct patterns in their similarity to the closest simulated lineage. For demonstration, we chose three tools, Freyja, Lollipop and VaQuERo, to represent three distinct patterns (Figure 2). Freyja’s false positives were generally highly similar to the closest true lineages, with similarity values concentrated near or above 0.9 in both child and parent categories and bimodal for lineages from the sibling clade category. Lollipop showed a broader distribution of similarities, with false positives spanning a wide range in all relationship categories and mean similarities centered around intermediate values. VaQuERo produced relatively few false-positive calls overall, and those that occurred tended to show high similarity to the closest simulated lineage, irrespective of the relationship.

**Figure 2:**
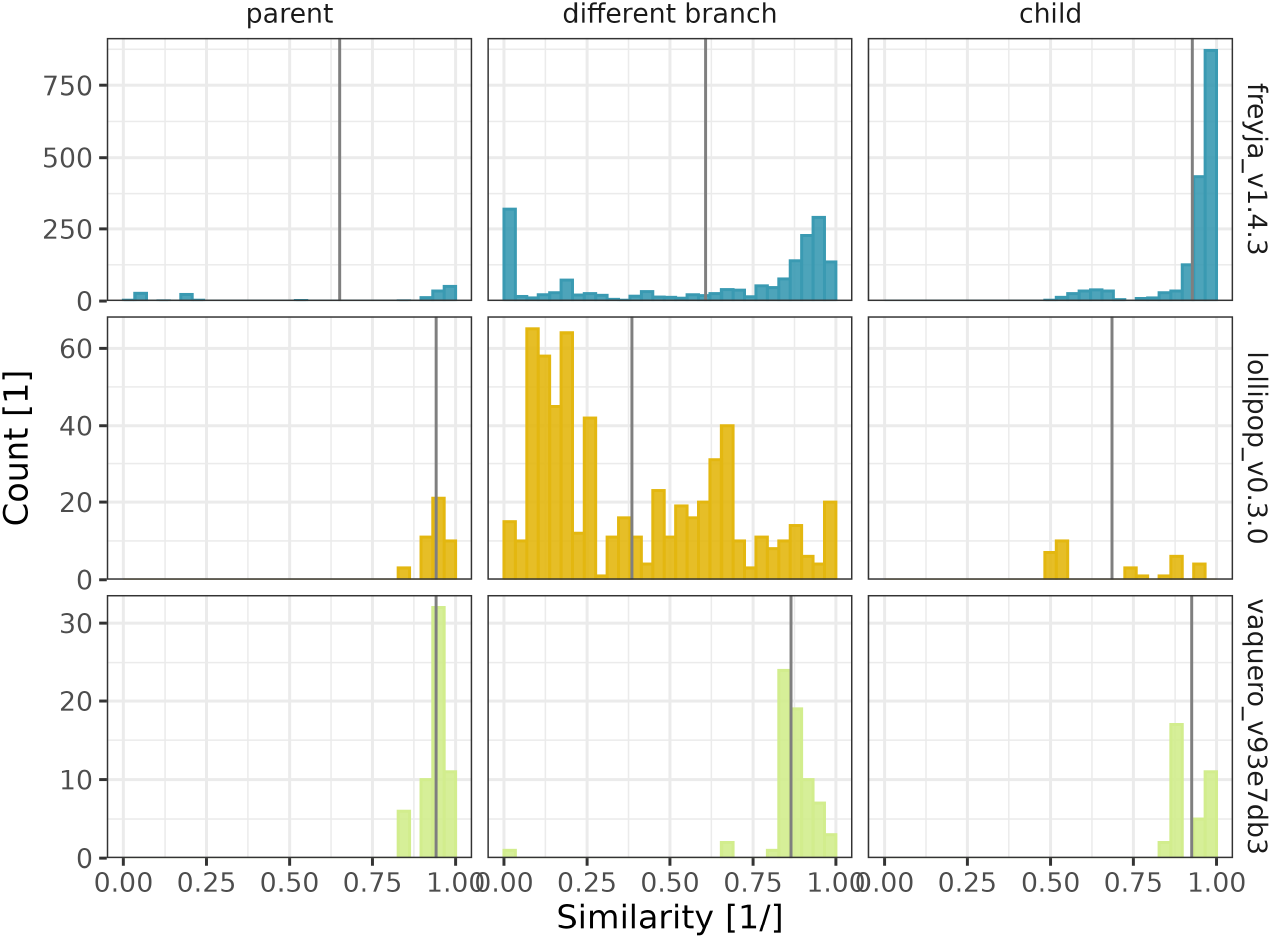
Distribution of false-positive lineage calls across three SARS-CoV-2 deconvolution tools (Freyja v1.4.3, Lollipop v0.3.0, and VaQuERo v93e7db3) aggregated across all simulated data sets. For each tool, histograms show the similarity between each false-positive lineage and its closest corresponding simulated lineage, with similarity ranging from 0 to 1 and defined as the proportion of shared mutations. False positives are grouped into three phylogenetic relationship categories: child (false positive represents a descendant of the closest simulated lineage), parent (false positive is an ancestor), and different branch (false positive is on an unrelated branch of the phylogeny). The darker vertical line in each panel represents the mean similarity within that category. The figure highlights differences in how each tool assigns incorrect lineages, both in the number of false positives and in their phylogenetic proximity to the closest true simulated lineages.

Overall, the false-positive patterns reveal clear differences in how each tool misassigns lineages. Partly, this can be explained by the different lineage resolutions provided by the reference sets used by the different tools (Table 1). This is accounted for in the lineage identification accuracy assessment by by considering a range of metrics that are independent of true negatives (see Supplemental File 1). Furthermore, different levels of ambiguity are allowed when determining true classifications. The ambiguity level defines how many sub-levels below a truly simulated lineage are still considered a true hit.

**Table 1:**
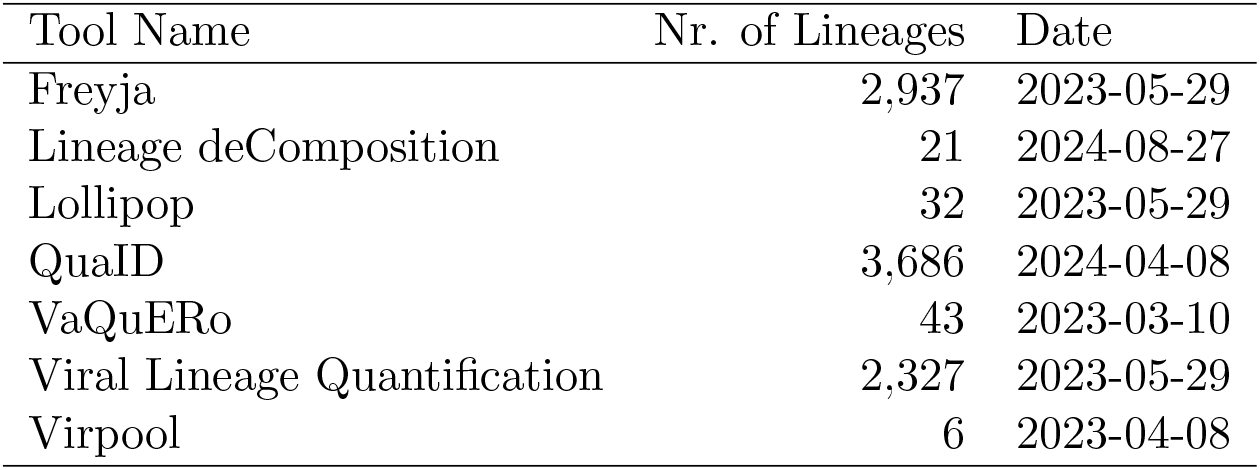
Scope and source of the tool specific lineage defining reference sets.

| Tool Name | Nr. of Lineages | Date |
| --- | --- | --- |
| Freyja | 2,937 | 2023-05-29 |
| Lineage deComposition | 21 | 2024-08-27 |
| Lollipop | 32 | 2023-05-29 |
| QuaID | 3,686 | 2024-04-08 |
| VaQuERo | 43 | 2023-03-10 |
| Viral Lineage Quantification | 2,327 | 2023-05-29 |
| Virpool | 6 | 2023-04-08 |

Depending on the scenario, such aggregation may mask important relationships between the identified and expected lineages. Even a single mutation can lead to an altered growth trajectory, as was the case during the emergence of the SARS-CoV-2 lineage JN.1 [39]. In such cases, the interpretation of a detection event, or the absence thereof, requires an understanding of the tendency of each tool to call fine-grained lineages conservatively or more permissively. Læmple aims to assess each tool’s behavior by contextualizing false positives with respect to the mutational distance and phylogenetic relationship to the closest variant included in the set of ground-truth variants. Freyja and VaQuERo tend to generate false positives that are closely related to the simulated lineages, with most errors occurring within child or parent relationships. This indicates that their misclassifications largely involve adjacent positions in the phylogeny, where lineages share substantial mutational overlap. VaQuERo shows particularly tight clustering in these categories, suggesting a strong tendency to confuse direct descendants or ancestors of the correct lineage rather than unrelated ones. In contrast, Lollipop produces a wider distribution of false positives, spanning both closely related and distantly related branches. Its false positives are more evenly spread across the child, parent, and different branch categories, and often exhibit lower similarity to the simulated lineages. Taken together, these results suggest that Freyja and VaQuERo primarily make proximate phylogenetic misclassifications, whereas Lollipop more frequently introduces false positives from remote parts of the lineage tree. Freyja tends to assign child lineages more often, whereas VaQuERo behaves more conservatively and tends to default to parent lineages.

### 6.3 Impact of sample complexity on tool performance

In our example workflow, we observed different performance characteristics, especially between Freyja and VaQuERo. Across all data sets, the two lineage-deconvolution tools showed distinct performance profiles with respect to PPV and TPR (Figure 3). In the VOC time-course, VaQuERo reached the highest precision, with a PPV of 1.0 and a TPR of about 0.7, whereas Freyja achieved a PPV of 0.5 and the same TPR. In the Omicron time-course, both tools showed reduced performance, but VaQuERo maintained higher PPV (0.5) compared to Freyja (~0.33), with reversed TPRs (VaQuERo 0.33 and Freyja 0.5). In the realistic Vienna time-course, Freyja showed a balanced performance with PPV and TPR both near 0.5, while VaQuERo displayed similar PPV (0.5) but markedly lower TPR (~0.15). When aggregating across all data sets, VaQuERo showed identical overall PPV (0.5), whereas Freyja exhibited higher overall TPR (0.5).

**Figure 3:**
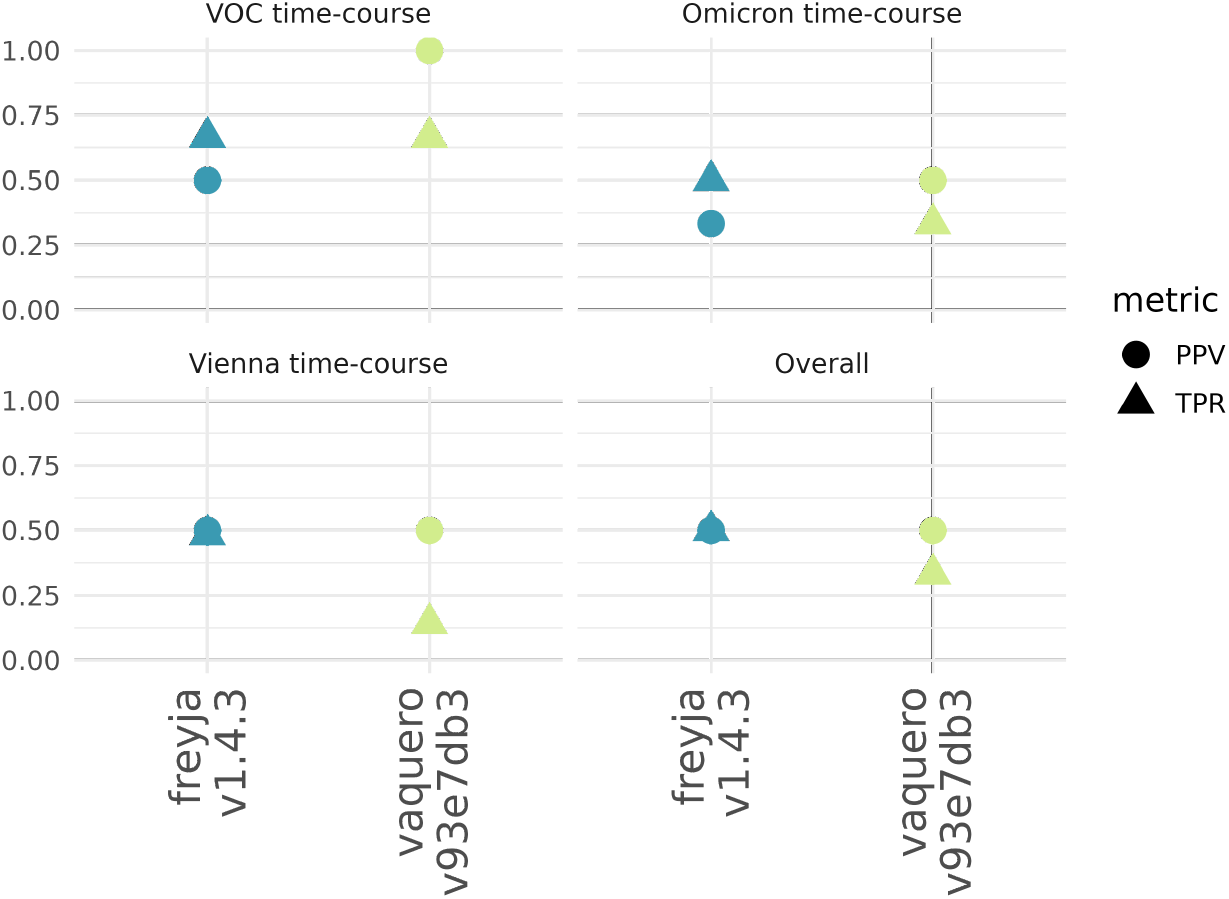
Comparison of SARS-CoV-2 lineage deconvolution performance between Freyja (v1.4.3) and VaQuERo (v93e7db3) across three simulated datasets (Omicron time-course, VOC timecourse, Vienna time-course) and an overall aggregated analysis. For each tool and dataset, two metrics are shown: positive predictive value (PPV; circles) and true positive rate (TPR; triangles). Higher PPV indicates better precision in avoiding false-positive lineage calls, while higher TPR indicates better sensitivity in detecting true lineages present in a sample. Points represent the median performance of each tool.

This comparison shows that VaQuERo achieves near-perfect precision and high sensitivity when lineages are highly divergent. In scenarios involving very closely related lineages, VaQuERo maintains higher precision than Freyja. In realistic, heterogeneous samples with many co-circulating lineages, Freyja identifies more true lineages, whereas VaQuERo remains more conservative, generating fewer false positives but missing more true lineages. Overall, VaQuERo exhibits a more precision-oriented profile, while Freyja shows a greater emphasis on sensitivity.

### 6.4 Impact of reference set on Sensitivity in VaQuERo

Using the same VaQuERo version (v24d9211) with two different reference sets resulted in noticeable differences in performance metrics (Figure 4). When run with the default reference set, VaQuERo reached a PPV of approximately 0.67 and a TPR of 0.16. In contrast, using the old reference set produced the same PPV (~0.67) but a substantially higher TPR (0.50). Thus, while precision remained stable across reference sets, sensitivity increased when the old reference set was used, which is in alignment with the expectation, since the old reference set provides more lineage resolution for lineages used in the simulated data sets, and at the same time contains fewer overall lineages, thereby simplifying the deconvolution task.

**Figure 4:**
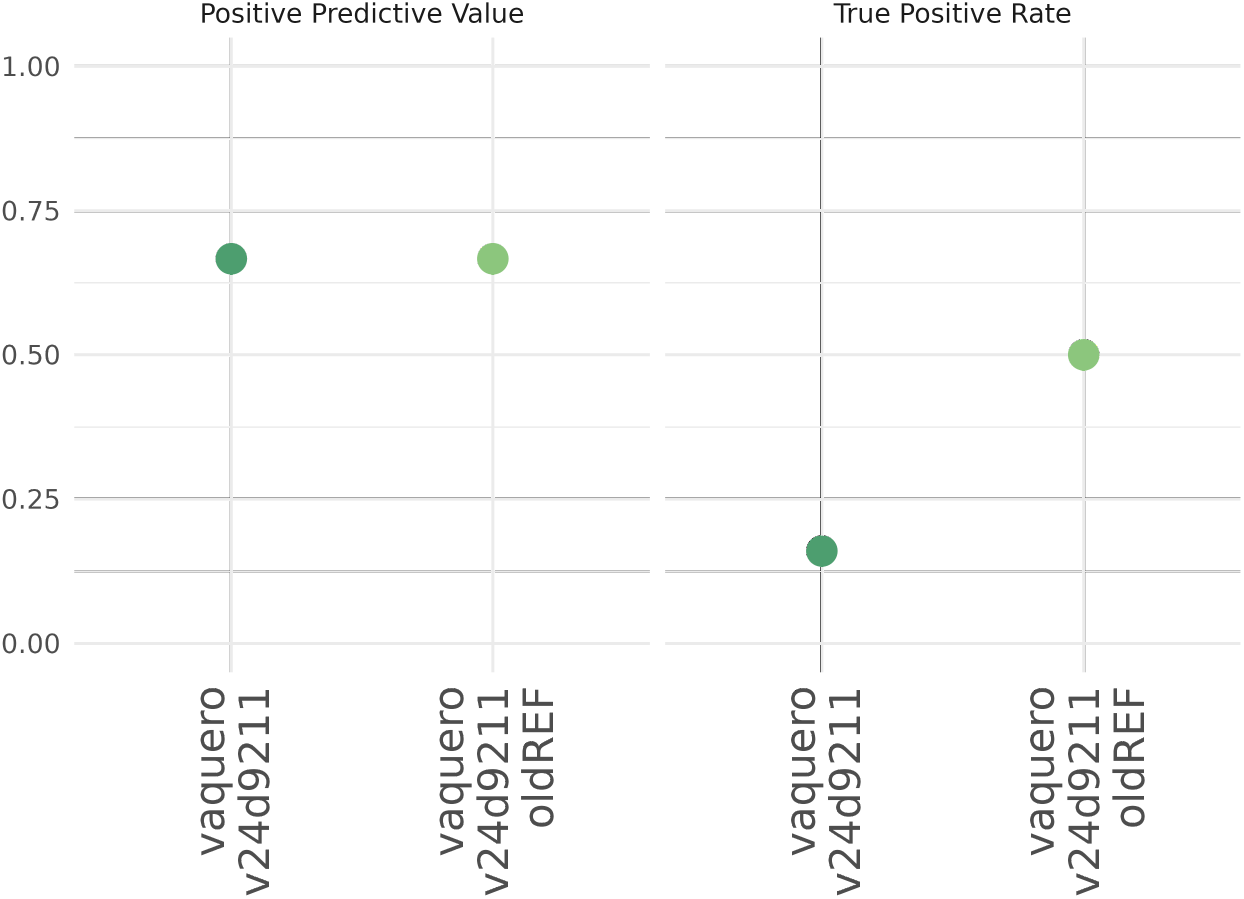
Comparison of lineage identification performance of the SARS-CoV-2 lineage deconvolution tool VaQuERo v24d9211 using either the default or an older reference data set (oldREF). Positive predictive values (PPV; left panel) and true positive rate values (TPR; right panel) are shown as individual data points for each reference configuration, representing the median value aggregated over all data sets. Higher PPV indicates greater precision in avoiding false-positive lineage calls, while higher TPR reflects improved sensitivity in detecting true lineages present in the sample.

### 6.5 Effect of Genome Coverage on Tool Precision

Precision or Positive Predictive Value (PPV) showed substantial variation across tools and sequencing coverage levels (Figure 5). Freyja, VirPool and Lollipop displayed moderate and relatively stable PPV values (0.3-0.45), with slight increases toward mid-range coverage before plateauing. VaQuERo showed low PPV at very low coverage but increased significantly with higher coverage, peaking around ~0.70 PPV at around 75 % genome coverage. QuaID and VLQ maintained low PPV throughout, with values generally below 0.30 across the entire coverage range. Overall, most tools showed only slight improvements in precision as coverage increased. These results indicate that higher sequencing coverage does not uniformly enhance PPV across deconvolution methods, and tool-specific behavior strongly influences precision outcomes.

**Figure 5:**
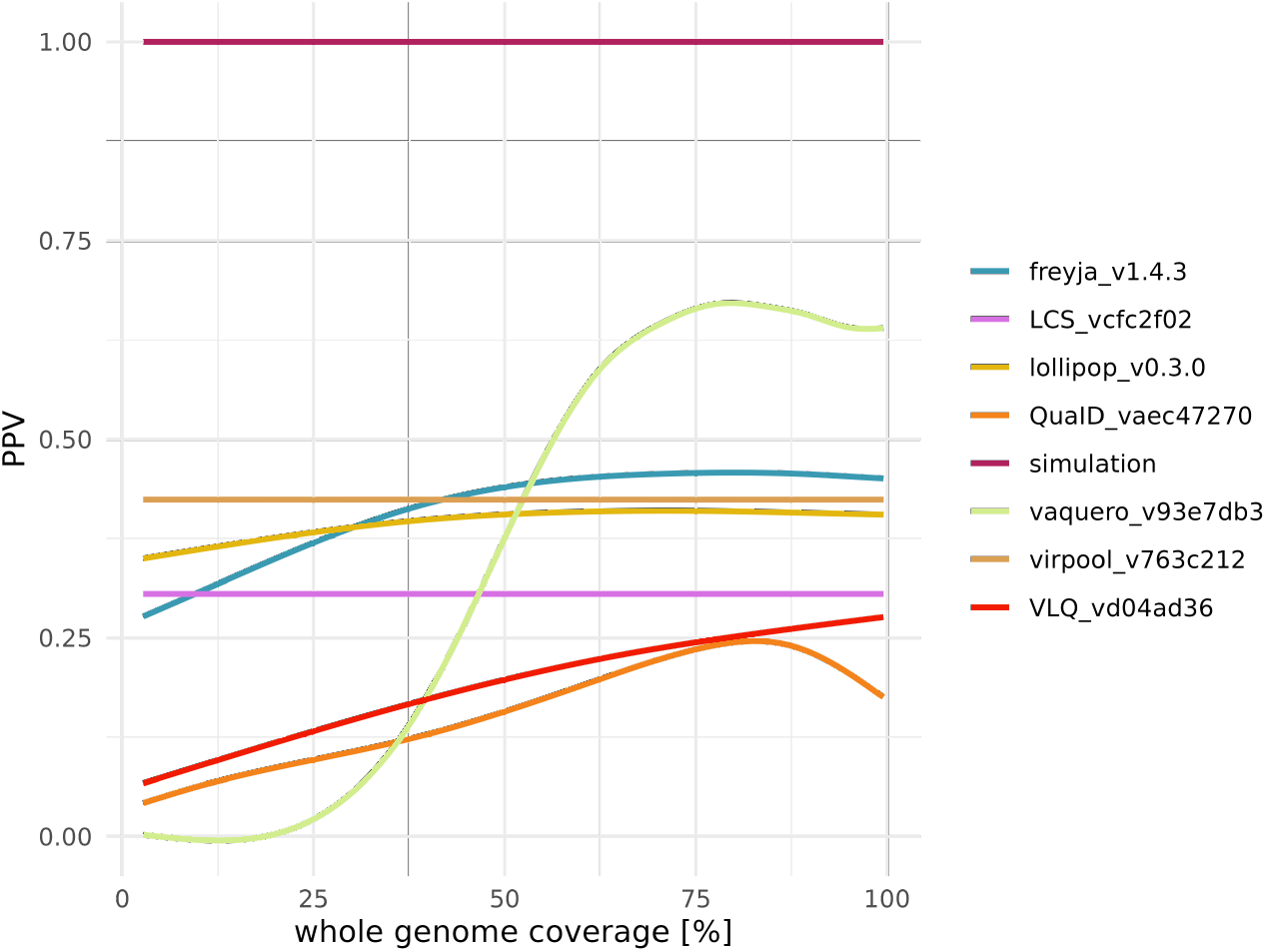
Positive predictive value (PPV) of multiple SARS-CoV-2 lineage deconvolution tools across a range of genome coverage levels over all tested data sets and plotted using a generalized additive model for smoothing. Higher PPV indicates greater precision in identifying only true lineages. A simulated benchmark is included that represents ideal performance. Most tools show only slight improvements in PPV as coverages increase, except for VaQuERo which exhibits a drastic increase in performance around 50 % genome coverage, illustrating tool-specific patterns of stability or sensitivity to coverage. conditions, the high-quality simulations appear representative of the sequencing quality achieved in our routine surveillance efforts.

### 6.6 Comparison of Simulated and Real-World Wastewater Data

To assess how well the simulated data reproduced characteristics of real-world wastewater sequencing data, we applied the evaluated deconvolution tools to real wastewater data from Vienna, collected during 2022, a period for which extensive clinical sequencing data were available to derive lineage ground truth [6]. The Læmple framework allows users to skip the simulation steps and instead process sequencing data and corresponding expected variant frequencies, if provided. Comparison of whole-genome coverage distributions showed that the simulated high-quality data closely resembled the coverage profiles observed in the real wastewater samples (Figure 6A). While the simulation framework additionally allows the generation of lower-quality data sets to evaluate tool robustness under adverse

**Figure 6:**
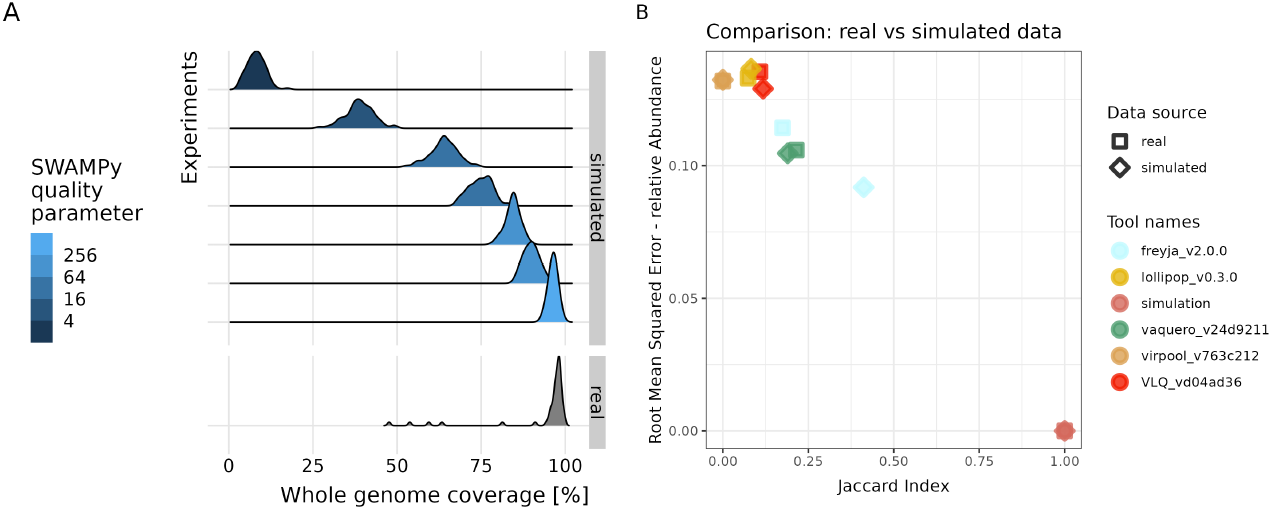
(A) Comparison of whole-genome coverage, as a proxy for sequencing quality, between simulated and real-world data sets. (B) Comparison of qualitative and quantitative lineage deconvolution performance obtained from real-world samples and from simulated samples of comparable sequencing quality, demonstrating that tool performance trends observed in simulated data closely resemble those observed in real-world wastewater samples.

Using clinical sequencing data from the same period and region, obtained through GISAID [37], as a reference, we calculated both the Jaccard Index for lineage identification and the Root Mean Squared Error (RMSE) for lineage abundance estimation in the real-world samples and in corresponding simulations. Overall, tool performance closely mirrored the patterns observed in the simulated benchmarking data (Figure 6B). Tools that achieved high lineage identification accuracy and abundance estimation performance in the simulation framework generally also performed well on the real-world data. In particular, the relative ranking of tools was largely preserved across both analyses. The main exception was Freyja, which showed somewhat stronger performance on simulated than on real-world samples. Taken together, these findings indicate that the simulation framework captures key properties of real wastewater sequencing data and produces benchmark results that are broadly predictive of real-world tool performance.

## 7 Discussion

Our demonstration analysis based on the example workflow revealed clear differences in how different deconvolution tools perform under varying levels of data set complexity. For example, tools such as Freyja and VaQuERo showed comparatively high sensitivity and precision respectively, while others showed greater variability across metrics and scenarios. Importantly, these performance patterns shifted substantially depending on lineage diversity, sequencing quality, and reference set composition. Rather than serving as definitive rankings, these results illustrate the broader point that tool performance is highly context-dependent and that systematic scenario-specific benchmarking is essential for meaningful interpretation of wastewater-based surveillance outputs.

The primary contribution of this work is therefore the benchmarking framework itself, which enabled the presented comparative findings. Wastewater-derived genomic data are inherently complex, shaped by mixed viral populations, convergent mutations, uneven sequencing coverage, and environmental noise. Our workflow addresses this complexity by providing a modular, reproducible, and scalable system for generating realistic in silico data sets, executing diverse deconvolution tools, and evaluating performance using standardized metrics. Because the entire process is implemented within a single Snakemake workflow, users can easily explore how tool performance responds to changes in lineage composition, mixture complexity, or sequencing characteristics, which are hard to capture comprehensively with static spike-in data sets.

In contrast to previous benchmarking studies relying on fixed mixtures of synthetic lineage controls, our approach offers a flexible and fully customizable alternative. While experimental spike-ins capture important laboratory-induced biases, they are costly, limited in the number of lineages they can include, and quickly become outdated as SARS-CoV-2 evolves. The ability to incorporate arbitrarily complex mixtures and newly emerging lineages into the simulation step allows the framework to assess tool performance as the viral population shifts, thereby assisting in quality control during tool maintenance and operation. This adaptability ensures that both tool developers and surveillance practitioners can evaluate performance under conditions that closely mirror current epidemiological challenges.

The framework also supports transparency and reproducibility, enabling direct comparison across software versions, parameter sets, or reference panels. This is particularly valuable given that many tools depend heavily on their reference sets, yet the relationship between reference curation and performance is rarely systematically examined [20]. By providing a consistent evaluation environment, the workflow helps to identify whether improvements stem from algorithmic changes or from updated reference data, and whether a tool’s strengths align with specific surveillance goals, such as early detection of emerging lineages or robust inference of trends in established variants.

Overall, the benchmarking workflow is designed as a living resource for the genomic surveillance community. Its modular nature ensures that new tools, simulation strategies, or evaluation metrics can be integrated with minimal effort, supporting continuous assessment as the methodological landscape evolves. Ultimately, this framework empowers researchers, public health laboratories, and tool developers to make informed, context-aware decisions when interpreting deconvolution results or selecting analytical pipelines. By emphasizing flexibility, realism, and reproducibility, it helps bridge the gap between methodological development and real-world surveillance needs.

## Data Availability

All data produced are available online on GitHub (https://github.com/Atotenschaedel/laemple_workflow)

## 8 Declarations

### 8.2 Authors’ Contributions

**Conceptualization**: AS, AB, FA; **Data Curation**: AS; **Formal Analysis**: AS, FA; **Methodology**: AS, FA; **Project Administration**: AB, FA; **Resources**: AB; **Software**: AS, FA; **Supervision**: AB, FA; **Validation**: AS; **Visualization**: AS, FA; **Writing – original draft**: AS, AB, FA; **Writing – review & editing**: AS, AB, FA;

## 9 Availability of Source Code and Requirements

The presented framework to assess the performance of virus lineage deconvolution tools for wastewater epidemiology, named Læmple, is freely available at GitHub (https://github.com/Atotenschaedel/laemple_workflow and free to use under the GNU General Public License (GPL). All data presented in this contribution can be reproduced therefrom.

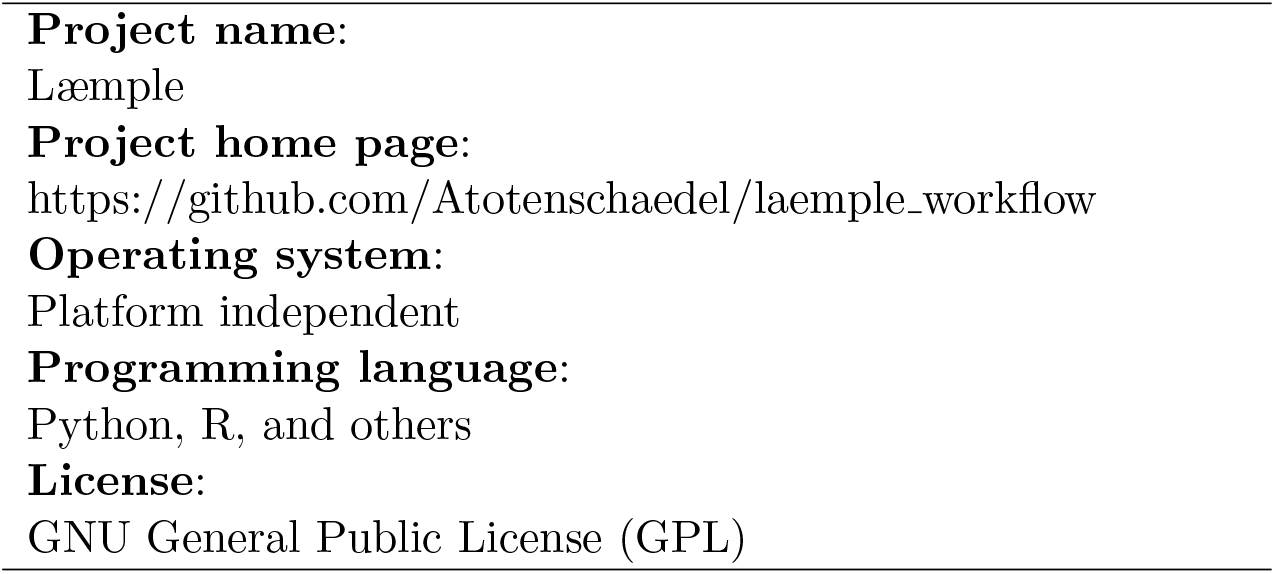

## 9.1 List of abbreviations

FNR: False Negative Rate
JI: Jaccard index
NGS: Next-generation sequencing
PPV: Positive Predictive Value
RMSE: Root Mean Squared Error
SARS-CoV-2: Severe Acute Respiratory Syndrome Coronavirus 2
TPR: True Positive Rate
VCF: Variant Call Format
VOC: Variant of Concern
WBS: Wastewater-based surveillance

