## Supplemental Material for "Læmple: A Benchmarking Framework for Virus Lineage Deconvolution Tools for SARS-CoV-2 from Wastewater"

### 1 Supplemental Files

#### 1.1 Supplemental File 1

Accompanying to this publication, a Supplemental File depicts the report as a HTML file. The report was generated with the Læmple framework and presents the results of the exemplary analysis, which forms the basis of the result section of the main text.

### 2 Supplemental Text

#### 2.1 Comments on user experience during installation and use

A total of seven deconvolution tools are being compared. Default parameters were used as often as possible to remove bias and if reference sets were provided, the most current, as of 2023-09-12 and broadest reference sets were chosen, unless otherwise stated.

##### 2.1.1 Lollipop

Lollipop [1] is installed via conda installation on 2023-05-29 (commitID: 3e57e5e). It requires an user-generated reference set for lineage calling. Following the installation guide from the public repository, lineage definitions were fetched from Public Health England (PHE) genomic repository (ukhsa-collaboration, 2023) and transformed into appropriate input format following the documentation provided. The reference set includes a total of 32 standardized lineage definition on the time of download.

#### 2.1.2 VaQuERo

VaQuERo [2] version 2 is installed from the GitHub repository (commitID: 08cc69c). Multiple reference sets are provided; therefore, the most recent ones were chosen. For the list of marker mutations, the used set was Europe, 2023-03-10, which includes a total of 43 lineages. Only one option was available for problematic mutation files and were therefore chosen as default. VaQuERo requires sample dates as input, therefore the first time-point of the simulation was given the date 2023-01-01 and all following time-points calculated accordingly in single day steps. For all other metadata the example data from the software documentation provided in the repository was used instead.

#### 2.1.3 Freyja

Freyja [3] was installed via software package manager conda on 2023- 05-29, based on GitHub repository version (commitID: 26b4063). The conda installation already includes a default reference set, only the reference genome for Wuhan-01 needs to be provided. The default reference set includes 2,937 lineages. Freyja was run using the variant calling file (VCF) from the variant calling pipeline described above using default parameters described in the installation guide from the public repository.

#### 2.1.4 Lineage deComposition

LCS [4] was installed on 2024-08-27 via GitHub repository (commitID: cfc2f02). The installation was straightforward and completely documented. Pre-generated marker source files are available and the most recent one, based on pango-designation (v.1.2.60) used for further analysis containing marker for 21 variants.

#### 2.1.5 QuaID

QuaID [5] was installed on 2024-04-08 from the public GitHub repository (commitID: aec47270). It requires installation of additional packages outside the repository (e.g., vdb), which further requires further installation of swift programming language for compiling, as well as additional python packages to generate the reference database. No default reference database was provided and was generated following the installation guide. Pre-generated MSA (multiple sequence alignment file) and Metadata file were downloaded from GISAID on 2024-11-25 and used to generate the reference database, which contains 3,686 variants. Default parameters were chosen as described in the publicly provided documentation. QuaID is returning results on the WHO Variant of Concern name level only, while all other tools in this study returned PANGO lineages. To make the results comparable, we opted to split all relative abundance estimates from QuaID over all pangolin lineages associated with each WHO variant of concern based on WHO historical variant of concern working definitions (outdated as of 15 March 2023). The historical working definitions were chosen as the WHO changed its working definitions for naming SARS-CoV-2 variants.

#### 2.1.6 Viral Lineage Quantification

VLQ [6] was installed on 2023-05-29 from the public GitHub repository (commitID: d04ad36), by installing all needed dependencies via conda installation channels as described by the documentation. Package versions were chosen according to the ones presented in the publication. VLQ requires a manually built reference set and does not provide a default reference set. Sequences and connected metadata were downloaded from the GISAID database. Only whole genome sequences from Europe were considered, however no further filter for region or sampling time was applied, which resulted in a total of 3,627 individual genome sequences. The further processing of the GISAID sequences was in accordance with the viral lineage quantification pipeline documentation, default filter parameters were chosen, however no further location or time period restriction was applied. The same SARS-CoV-2Wuhan-01 as for the other tools was chosen for reference SARS-CoV-2 genome sequence. Eventually, the reference set includes 2,327 lineages.

#### 2.1.7 VirPool

VirPool [7] was installed on 2023-04-08 from the public GitHub repository (commit: 763c212). Installation via conda environment command using provided required package list was fast and only took a few minutes. A test case was provided and up-to-date. Multiple pre-generated variant profiles are provided. Variant profile for Austria containing 6 variants was chosen as the most appropriate for simulated test cases, as one of the simulated time courses is based on data from Vienna.

[3] Smruthi Karthikeyan, Joshua I. Levy, Peter De Hoff, Greg Humphrey, Amanda Birmingham, Kristen Jepsen, Sawyer Farmer, Helena M. Tubb, Tommy Valles, Caitlin E. Tribelhorn, Rebecca Tsai, Stefan Aigner, Shashank Sathe, Niema Moshiri, Benjamin Henson, Adam M. Mark, Abbas Hakim, Nathan A. Baer, Tom Barber, Pedro Belda-Ferre, Marisol Chacón, Willi Cheung, Evelyn S. Cresini, Emily R. Eisner, Alma L. Lastrella, Elijah S. Lawrence, Clarisse A. Marotz, Toan T. Ngo, Tyler Ostrander, Ashley Plascencia, Rodolfo A. Salido, Phoebe Seaver, Elizabeth W. Smoot, Daniel McDonald, Robert M. Neuhard, Angela L. Scioscia, Alysson M. Satterlund, Elizabeth H. Simmons, Dismas B. Abelman, David Brenner, Judith C. Bruner, Anne Buckley, Michael Ellison, Jeffrey Gattas, Steven L. Gonias, Matt Hale, Faith Hawkins, Lydia Ikeda, Hemlata Jhaveri, Ted Johnson, Vince Kellen, Brendan Kremer, Gary Matthews, Ronald W. McLawhon, Pierre Ouillet, Daniel Park, Allorah Pradenas, Sharon Reed, Lindsay Riggs, Alison Sanders, Bradley Sollenberger, Angela Song, Benjamin White, Terri Winbush, Christine M. Aceves, Catelyn Anderson, Karthik Gangavarapu, Emory Hufbauer, Ezra Kurzban, Justin Lee, Nathaniel L. Matteson, Edyth Parker, Sarah A. Perkins, Karthik S. Ramesh, Refugio Robles-Sikisaka, Madison A. Schwab, Emily Spencer, Shirlee Wohl, Laura Nicholson, Ian H. Mchardy, David P. Dimmock, Charlotte A. Hobbs, Omid Bakhtar, Aaron Harding, Art Mendoza, Alexandre Bolze, David Becker, Elizabeth T. Cirulli, Magnus Isaksson, Kelly M. Schiabor Barrett, Nicole L. Washington, John D. Malone, Ashleigh Murphy Schafer, Nikos Gurfield, Sarah Stous, Rebecca Fielding-Miller, Richard S. Garfein, Tommi Gaines, Cheryl Anderson, Natasha K. Martin, Robert Schooley, Brett Austin, Duncan R. MacCannell, Stephen F. Kingsmore, William Lee, Seema Shah, Eric McDonald, Alexander T. Yu, Mark Zeller, Kathleen M. Fisch, Christopher Longhurst, Patty Maysent, David Pride, Pradeep K. Khosla, Louise C. Laurent, Gene W. Yeo, Kristian G. Andersen, and Rob Knight. Wastewater sequencing reveals early cryptic SARS-CoV-2 variant transmission. *Nature*, pages 1–4, 7 2022. ISSN 1476-4687. doi: 10.1038/s41586-022-05049-6. publisher: Nature Publishing Group.

[4] Renan Valieris, Rodrigo D. Drummond, Alexandre Defelicibus, Emmanuel Dias-Neto, Rafael A. Rosales, and Israel Tojal da Silva. A mixture model for determining SARS-Cov-2 variant composition in pooled samples. *Bioinformatics (Oxford, England)*, 38(7):1809–1815, 3 2022. ISSN 1367-4811. doi: 10.1093/bioinformatics/btac047. PMID: 35104309.

[5] Nicolae Sapoval, Yunxi Liu, Esther G. Lou, Loren Hopkins, Katherine B. Ensor, Rebecca Schneider, Lauren B. Stadler, and Todd J. Treangen. Enabling accurate and early detection of recently emerged SARS-CoV-2 variants of concern in wastewater. *Nature Communications*, 14(1):2834, 5 2023. ISSN 2041-1723. doi: 10.1038/s41467-023-38184-3. number: 1 publisher: Nature Publishing Group.

[6] Jasmijn A. Baaijens, Alessandro Zulli, Isabel M. Ott, Ioanna Nika, Mart J. van der Lugt, Mary E. Petrone, Tara Alpert, Joseph R. Fauver, Chaney C. Kalinich, Chantal B. F. Vogels, Mallery I. Breban, Claire Duvallet, Kyle A. McElroy, Newsha Ghaeli, Maxim Imakaev, Malaika F. Mckenzie-Bennett, Keith Robison, Alex Plocik, Rebecca Schilling, Martha Pierson, Rebecca Littlefield, Michelle L. Spencer, Birgitte B. Simen, Ahmad Altajar, Anderson F. Brito, Anne E. Watkins, Anthony Muyombwe, Caleb Neal, Chen Liu, Christopher Castaldi, Claire Pearson, David R. Peaper, Eva Laszlo, Irina R. Tikhonova, Jafar Razeq, Jessica E. Rothman, Jianhui Wang, Kaya Bilguvar, Linda Niccolai, Madeline S. Wilson, Margaret L. Anderson, Marie L. Landry, Mark D. Adams, Pei Hui, Randy Downing, Rebecca Earnest, Shrikant Mane, Steven Murphy, William P. Hanage,

Nathan D. Grubaugh, Jordan Peccia, Michael Baym, and Yale SARS-CoV-2 Genomic Surveillance Initiative. Lineage abundance estimation for SARS-CoV-2 in wastewater using transcriptome quantification techniques. *Genome Biology*, 23(1):236, 11 2022. ISSN 1474-760X. doi: 10.1186/s13059-022-02805-9.

[7] Askar Gafurov, Andrej Baláž, Fabian Amman, Kristína Boršová, Viktória Čabanová, Boris Klempa, Andreas Bergthaler, Tomáš Vinař, and Broňa Brejová. Virpool: model-based estimation of SARS-CoV-2 variant proportions in wastewater samples. *BMC Bioinformatics*, 23(1):551, 12 2022. ISSN 1471-2105. doi: 10.1186/s12859-022-05100-3.
