## Supplemental File 1 for "Læmple: A Benchmarking Framework for Virus Lineage Deconvolution Tools for SARS-CoV-2 from Wastewater": Supplemental_File_1.html

Læmple: Virus Lineage Deconvolution Benchmarking


Code 

- Show All Code
- Hide All Code

### Læmple: Virus Lineage Deconvolution Benchmarking

##### Comprehensive Report

###### Anna Schedl

#### 2026-07-21 18:57:20.678093

```
# Read config file from params
config <- yaml::read_yaml(params$config_file)

# color for simulation data
tool_list <- c("simulation"="#D5695DFF")

# additional labels
label.legend <- c("simulation" = "simulation",
                  "P"="Real Positive", "N"="Real Negative", 
                  "PP"="Predicted Positive", "TP"="True Positive", "FP"="False Positive", "FN" = "False Negative", 
                  "RMSE"="Root Mean Square Error", 
                  "TPR"="True Positive Rate", 
                  "FNR"="False Negative Rate", "PPV"="Positive Predictive Value", "FDR"="False Discovery Rate", 
                  "jaccard_index"="Jaccard Index", "F1_score"="F1 score", "count_lineage"="Number of lineages", 
                  "uniformity_wg_per"="Percent, genome min coverage",
                  "different branch" = "different branch", "parent"="parent", "child"="child",
                  "Overall" = "Overall"
                  )

for (label in config$POSTPRED$LABELS){
  label.legend[label$key] <- label$value
}

for (tool in config$TOOLS){
  if (tool$INCLUDE_IN_ANALYSIS){
    tool_list[tool$TOOL_NAME] <- tool$COLOUR_IN_REPORT
    label.legend[tool$TOOL_NAME] <- tool$TOOL_LABEL
  }
}

# safe_labeller function, returns original value if not found in label.legends
safe_labeller <- function(x) {
  if (is.data.frame(x)) {
    out <- lapply(x, function(col) {
      mapped <- label.legend[as.character(col)]
      mapped[is.na(mapped)] <- as.character(col)[is.na(mapped)]
      mapped
    })
    return(as.data.frame(out, stringsAsFactors = FALSE))
  }

  mapped <- label.legend[as.character(x)]
  mapped[is.na(mapped)] <- as.character(x)[is.na(mapped)]
  mapped
}

#set colors
colScale <- scale_colour_manual(values = tool_list)
fillScale <- scale_fill_manual(values=tool_list)

#set up json data
url <- "https://raw.githubusercontent.com/corneliusroemer/pango-sequences/refs/heads/main/data/pango-consensus-sequences_summary.json"
file = paste(currWorkingDir, "/reference/consensus_sequences/data/pango-consensus-sequences_summary.json", sep="")
json.data <- jsonlite::fromJSON(url)

#clean up
rm(label, tool)
```

### Introduction

Graphical overview of the benchmarking results of the current Læmple
analysis.

Læmple simulates a time course with a sequence of changing virus
lineage abundances, from which a predefined number of time points are
samples and used to simulate whole genome tiling amplicon sequencing
data, using a third party tool named SWAMPy. The
sequencing quality can vary between different experiments, and be used
as an additional covariate of the analysis. Each experiment is performed
in a predefined number of replicas.

On these simulated reads virus variant deconvolution tools are
applied.

In the following report, the results of these tools –with each other,
and to the simulated ground truth– are compared. Thereby qualitative
measures (e.g. False Negative Rate, Positive Predictive Value, Jaccard
Index) and quantitative measures (e.g. rooted mean squared error of
relative abundance) are deduced to assess individual tool performance.
Additionally, the observed errors, in particular false positive
classifications, are further evaluated with respect to the closest true
positive, to assess the severity of mis-classification calls.

---

#### Settings & Sample Overview

List included Tools:

```
knitr::kable(data.frame(Tools = names(tool_list)) %>% rowwise() %>% mutate(`Tool name` = unlist(str_split(Tools, "_"))[1], Version = unlist(str_split(Tools, "_"))[2], Reference = unlist(str_split(Tools, "_"))[3]) %>% dplyr::select(-Tools) %>% mutate(Reference = ifelse(is.na(Reference), "default", Reference)))
```

| Tool name | Version | Reference |
| --- | --- | --- |
| simulation | NA | default |
| freyja | v1.4.3 | default |
| freyja | v1.5.3 | default |
| freyja | v2.0.0 | default |
| LCS | vcfc2f02 | default |
| lollipop | v0.3.0 | default |
| QuaID | vaec47270 | default |
| vaquero | v24d9211 | default |
| vaquero | v24d9211 | oldREF |
| vaquero | v93e7db3 | default |
| virpool | v763c212 | default |
| VLQ | vd04ad36 | default |

---

```
experiment_list <- list.dirs(path=paste0(currWorkingDir, "/experiments"), full.names=FALSE, recursive=FALSE)
experiment_list <- experiment_list[grepl("Ex", experiment_list)]
experiment_list <- experiment_list[!grepl("^##", experiment_list)]
```

**Analysis in numbers:**

- Number of time courses considered: 3
- Number of time point(s) considered: 46
- Number of tool(s) considered: 11
- Number of experiment(s) considered: 21
- Number of replicate(s) considered: 3

```
data.metrics <- calculateConfusionMatrix(data.tool.long, data.sim.long)
```

**Calculate metrics for different ambiguity level
modes**

For the calculation of true classification different levels of
ambiguity are allowed – – The ambiguity level defines how many
sub-levels of a truely simulated lineage is still considered a true hit.
E.g., at level 1 a detection of “B.1.1” would still be a true
classification if “B.1” was in the set of simulated lineages. But
lineage “B”, or lineage “B.1.1.1” would not. The latter would still be a
true hit at the ambiguity level 2.

### Simulated Data

#### Simulated Lineage Abundance

Visualisation of the simulated relative lineage abundances per time
course.

##### VOC time-course

##### Omicron time-course

##### Vienna time-course

#### Simulated Coverage

Distribution of genome coverage for each of the simulated
experiments, as a proxy for overall sequencing quality.

##### VOC time-course

##### Omicron time-course

##### Vienna time-course

---

### Results

#### Comparing replicates

##### Venn Diagrams

Comparing the proportion of shared true classifications between the
replicas of the same time course.

###### VOC time-course

###### Omicron time-course

###### Vienna time-course

###### Overall

##### Density Plots

Comparing the distribution of performance metrics between the
replicas of the same time course.

###### VOC time-course

###### freyja\_v1.4.3

###### freyja\_v1.5.3

###### freyja\_v2.0.0

###### LCS\_vcfc2f02

###### lollipop\_v0.3.0

###### QuaID\_vaec47270

###### vaquero\_v24d9211

###### vaquero\_v24d9211\_oldREF

###### vaquero\_v93e7db3

###### virpool\_v763c212

###### VLQ\_vd04ad36

###### Omicron time-course

###### freyja\_v1.4.3

###### freyja\_v1.5.3

###### freyja\_v2.0.0

###### LCS\_vcfc2f02

###### lollipop\_v0.3.0

###### QuaID\_vaec47270

###### vaquero\_v24d9211

###### vaquero\_v24d9211\_oldREF

###### vaquero\_v93e7db3

###### virpool\_v763c212

###### VLQ\_vd04ad36

###### Vienna time-course

###### freyja\_v1.4.3

###### freyja\_v1.5.3

###### freyja\_v2.0.0

###### LCS\_vcfc2f02

###### lollipop\_v0.3.0

###### QuaID\_vaec47270

###### vaquero\_v24d9211

###### vaquero\_v24d9211\_oldREF

###### vaquero\_v93e7db3

###### virpool\_v763c212

###### VLQ\_vd04ad36

###### Overall

###### freyja\_v1.4.3

###### freyja\_v1.5.3

###### freyja\_v2.0.0

###### LCS\_vcfc2f02

###### lollipop\_v0.3.0

###### QuaID\_vaec47270

###### vaquero\_v24d9211

###### vaquero\_v24d9211\_oldREF

###### vaquero\_v93e7db3

###### virpool\_v763c212

###### VLQ\_vd04ad36

---

#### Compare experiments w/ different quality settings

Comparing the distribution of performance metrics between experiments
w/ different quality settings.

##### VOC time-course

##### Omicron time-course

##### Vienna time-course

---

#### Predicted timecoures

Predicted lineages and their abundance and comparision with true
expected lineage abundance given the underlying simulations.

##### VOC time-course

###### freyja\_v1.4.3

[1] “freyja\_v1.4.3” [1] “Errorbars are standard error over all
experiments. Showing top 36 lineage of 150 lineages (top lineage
according to mean of abundance over all timepoints and predicted at more
then 1 timepoint.) 1 false negative signals.”

###### freyja\_v1.5.3

[1] “freyja\_v1.5.3” [1] “Errorbars are standard error over all
experiments. Showing top 36 lineage of 166 lineages (top lineage
according to mean of abundance over all timepoints and predicted at more
then 1 timepoint.) 1 false negative signals.”

###### freyja\_v2.0.0

[1] “freyja\_v2.0.0” [1] “Errorbars are standard error over all
experiments. Showing top 36 lineage of 166 lineages (top lineage
according to mean of abundance over all timepoints and predicted at more
then 1 timepoint.) 1 false negative signals.”

###### LCS\_vcfc2f02

[1] “LCS\_vcfc2f02” [1] “Errorbars are standard error over all
experiments.”

###### lollipop\_v0.3.0

[1] “lollipop\_v0.3.0” [1] “Errorbars are standard error over all
experiments. Showing top 36 lineage of 37 lineages (top lineage
according to mean of abundance over all timepoints and predicted at more
then 1 timepoint.) 1 false negative signals.”

###### QuaID\_vaec47270

[1] “QuaID\_vaec47270” [1] “Errorbars are standard error over all
experiments.”

###### vaquero\_v24d9211

[1] “vaquero\_v24d9211” [1] “Errorbars are standard error over all
experiments.”

###### vaquero\_v24d9211\_oldREF

[1] “vaquero\_v24d9211\_oldREF” [1] “Errorbars are standard error over
all experiments.”

###### vaquero\_v93e7db3

[1] “vaquero\_v93e7db3” [1] “Errorbars are standard error over all
experiments.”

###### virpool\_v763c212

[1] “virpool\_v763c212” [1] “Errorbars are standard error over all
experiments.”

###### VLQ\_vd04ad36

[1] “VLQ\_vd04ad36” [1] “Errorbars are standard error over all
experiments. Showing top 36 lineage of 500 lineages (top lineage
according to mean of abundance over all timepoints and predicted at more
then 1 timepoint.)”

##### Omicron time-course

###### freyja\_v1.4.3

[1] “freyja\_v1.4.3” [1] “Errorbars are standard error over all
experiments. Showing top 36 lineage of 223 lineages (top lineage
according to mean of abundance over all timepoints and predicted at more
then 1 timepoint.) 2 false negative signals.”

###### freyja\_v1.5.3

[1] “freyja\_v1.5.3” [1] “Errorbars are standard error over all
experiments. Showing top 36 lineage of 327 lineages (top lineage
according to mean of abundance over all timepoints and predicted at more
then 1 timepoint.) 1 false negative signals.”

###### freyja\_v2.0.0

[1] “freyja\_v2.0.0” [1] “Errorbars are standard error over all
experiments. Showing top 36 lineage of 327 lineages (top lineage
according to mean of abundance over all timepoints and predicted at more
then 1 timepoint.) 1 false negative signals.”

###### LCS\_vcfc2f02

[1] “LCS\_vcfc2f02” [1] “Errorbars are standard error over all
experiments.”

###### lollipop\_v0.3.0

[1] “lollipop\_v0.3.0” [1] “Errorbars are standard error over all
experiments.”

###### QuaID\_vaec47270

[1] “QuaID\_vaec47270” [1] “Errorbars are standard error over all
experiments.”

###### vaquero\_v24d9211

[1] “vaquero\_v24d9211” [1] “Errorbars are standard error over all
experiments.”

###### vaquero\_v24d9211\_oldREF

[1] “vaquero\_v24d9211\_oldREF” [1] “Errorbars are standard error over
all experiments.”

###### vaquero\_v93e7db3

[1] “vaquero\_v93e7db3” [1] “Errorbars are standard error over all
experiments.”

###### virpool\_v763c212

[1] “virpool\_v763c212” [1] “Errorbars are standard error over all
experiments.”

###### VLQ\_vd04ad36

[1] “VLQ\_vd04ad36” [1] “Errorbars are standard error over all
experiments. Showing top 36 lineage of 502 lineages (top lineage
according to mean of abundance over all timepoints and predicted at more
then 1 timepoint.) 1 false negative signals.”

##### Vienna time-course

###### freyja\_v1.4.3

[1] “freyja\_v1.4.3” [1] “Errorbars are standard error over all
experiments. 22 lineages only simulated in 1 Timepoint and removed from
visualisation. Showing top 36 lineage of 436 lineages (top lineage
according to mean of abundance over all timepoints and predicted at more
then 1 timepoint.) 59 false negative signals.”

###### freyja\_v1.5.3

[1] “freyja\_v1.5.3” [1] “Errorbars are standard error over all
experiments. 22 lineages only simulated in 1 Timepoint and removed from
visualisation. Showing top 36 lineage of 435 lineages (top lineage
according to mean of abundance over all timepoints and predicted at more
then 1 timepoint.) 61 false negative signals.”

###### freyja\_v2.0.0

[1] “freyja\_v2.0.0” [1] “Errorbars are standard error over all
experiments. 22 lineages only simulated in 1 Timepoint and removed from
visualisation. Showing top 36 lineage of 435 lineages (top lineage
according to mean of abundance over all timepoints and predicted at more
then 1 timepoint.) 61 false negative signals.”

###### LCS\_vcfc2f02

[1] “LCS\_vcfc2f02” [1] “Errorbars are standard error over all
experiments. 22 lineages only simulated in 1 Timepoint and removed from
visualisation. Showing top 21 lineage of 21 lineages (top lineage
according to mean of abundance over all timepoints and predicted at more
then 1 timepoint.) 69 false negative signals.”

###### lollipop\_v0.3.0

[1] “lollipop\_v0.3.0” [1] “Errorbars are standard error over all
experiments. 22 lineages only simulated in 1 Timepoint and removed from
visualisation. Showing top 36 lineage of 36 lineages (top lineage
according to mean of abundance over all timepoints and predicted at more
then 1 timepoint.) 60 false negative signals.”

###### QuaID\_vaec47270

[1] “QuaID\_vaec47270” [1] “Errorbars are standard error over all
experiments. 22 lineages only simulated in 1 Timepoint and removed from
visualisation. Showing top 4 lineage of 4 lineages (top lineage
according to mean of abundance over all timepoints and predicted at more
then 1 timepoint.) 68 false negative signals.”

###### vaquero\_v24d9211

[1] “vaquero\_v24d9211” [1] “Errorbars are standard error over all
experiments. 22 lineages only simulated in 1 Timepoint and removed from
visualisation. Showing top 21 lineage of 21 lineages (top lineage
according to mean of abundance over all timepoints and predicted at more
then 1 timepoint.) 54 false negative signals.”

###### vaquero\_v24d9211\_oldREF

[1] “vaquero\_v24d9211\_oldREF” [1] “Errorbars are standard error over
all experiments. 22 lineages only simulated in 1 Timepoint and removed
from visualisation. Showing top 18 lineage of 18 lineages (top lineage
according to mean of abundance over all timepoints and predicted at more
then 1 timepoint.) 57 false negative signals.”

###### vaquero\_v93e7db3

[1] “vaquero\_v93e7db3” [1] “Errorbars are standard error over all
experiments. 22 lineages only simulated in 1 Timepoint and removed from
visualisation. Showing top 20 lineage of 20 lineages (top lineage
according to mean of abundance over all timepoints and predicted at more
then 1 timepoint.) 56 false negative signals.”

###### virpool\_v763c212

[1] “virpool\_v763c212” [1] “Errorbars are standard error over all
experiments. 22 lineages only simulated in 1 Timepoint and removed from
visualisation. Showing top 6 lineage of 6 lineages (top lineage
according to mean of abundance over all timepoints and predicted at more
then 1 timepoint.) 70 false negative signals.”

###### VLQ\_vd04ad36

[1] “VLQ\_vd04ad36” [1] “Errorbars are standard error over all
experiments. 22 lineages only simulated in 1 Timepoint and removed from
visualisation. Showing top 36 lineage of 690 lineages (top lineage
according to mean of abundance over all timepoints and predicted at more
then 1 timepoint.) 69 false negative signals.”

---

#### Lineage identifcation

Qualiative performance metrics for different time courses and
different ambiguity levels.

##### VOC time-course

###### Strict mode

Metrics - Strict
mode

###### Adj. 1 level

Metrics - WideQual - Adj.
1

###### Adj. 2 level

Metrics - WideQual - Adj.
2

##### Omicron time-course

###### Strict mode

Metrics - Strict
mode

###### Adj. 1 level

Metrics - nearQual - Adj.
1

###### Adj. 2 level

Metrics - nearQual - Adj.
2

##### Vienna time-course

###### Strict mode

Metrics - Strict
mode

###### Adj. 1 level

Metrics - realTimecourse - Adj.
1

###### Adj. 2 level

Metrics - realTimecourse - Adj.
2

##### Overall

###### Strict mode

Metrics - Overall - Strict
mode

###### Adj. 1 level

Metrics - Overall - Adj.
1

###### Adj. 2 level

Metrics - Overall - Adj.
2

---

#### Library complexity and Sequencing quality

##### Number of lineages per sample

Key quality measures as a function of the number of simulated
lineages.

##### Genomic coverage

Key quality measures as a function of the simulated sequencing
quality.

##### Abundance Estimation

Estimated lineage abundance as a function of simulated sequencing
quality and simulated lineage abundance.

##### Error Estimation

Residual of estimated lineage abundance as a function of simulated
lineage abundance.

---

#### False Positive - Phylogenetic Relationship

A characterisaton of all detected false positive lineages and their
phylogenic relation and similiarity (based on shared mutations) to the
closest true positive. Vertical line indicate the mean similarity for
each tool and group.

##### VOC time-course

##### Omicron time-course

##### Vienna time-course

##### Overall

---

### Summary: Jaccard index vs RMSE

Aggregation plot summarizing the qualitative classification
performance (Jaccard Index) and the quantitative inferrence performance
(RMSE: root mean squared error) for each tool.

#### VOC time-course

#### Omicron time-course

#### Vienna time-course

#### Overall
